# RT qLAMP--Direct Detection of SARS-CoV-2 in Raw Sewage

**DOI:** 10.1101/2020.10.01.20205492

**Authors:** Jerry E. Ongerth, Richard E. Danielson

## Abstract

The project purpose was to examine ability of a loop-mediated isothermal amplification (LAMP) assay to quantify SARS-CoV-2 in raw sewage, directly, using no preliminary sample processing for virus concentration and RNA extraction. An objective was to take advantage of extensive recently published work to facilitate process development, principally primer selection, and to use readily available off the shelf materials with conventional lab procedure and equipment. Well-developed and referenced primers for ORF1a, E, and N-gene targets were selected and applied, using commercially available synthetic RNA standards, and raw sewage from a local wastewater agency serving 650,000. County health department monitoring provided current COVID-19 data. Testing defined performance characteristics for each primer set, with significant differences between them. Specific amplification of SARS-CoV-2 RNA was observed using each of the primer sets, with E-gene and N-gene primers most effective. Positive analysis results from all raw sewage samples corresponded to calculated concentrations of virus in 5-10 μL raw sewage aliquots for 25 μL reactions. Results show that even at low reported case rates e.g. 1-10/100,000, SARS-CoV-2 is present in raw sewage at > 1-5/ μL, permitting direct LAMP-based detection. Use of RT qLAMP will facilitate wastewater-based epidemiology as an important component for COVID-19 control.

## Introduction

The purpose of this report is to describe the ability of loop-mediated isothermal amplification, LAMP, in the form of RT qLAMP to detect and quantify SARS-CoV-2 in raw sewage, directly…i.e. without sample processing for virus concentration or RNA extraction. We provide information on the routine application, equipment, and facilities used to illustrate the feasibility of RT qLAMP application for detailed monitoring of SARS-CoV-2 for wastewater based epidemiology (WBE). The most important and novel aspect of this report is demonstration that even at low reported case rates e.g. 1-10/100,000, in a community, SARS-CoV-2 virus is present in raw sewage at concentrations > 1-5/μL, sufficient for LAMP-based detection directly avoiding the qPCR need for cumbersome time-consuming concentration and RNA extraction. Incorporation of this analytical approach will facilitate development of data supporting wastewater-based epidemiology as an important component of policy advice directed to COVID-19 control.

Loop-mediated isothermal amplification, LAMP, is not novel and it is not new. It is a thoroughly demonstrated and well-understood nucleic acid amplification procedure, first described 20 years ago, Notomi, 2000, since developed for largely clinical applications but equally demonstrated for detection of DNA or RNA in a wide variety of viral, microbial, and protozoan pathogens as well as identifying gene-specific targets in plants and animals, e.g., Salar et al, 2013; Becherer et al, 2020. The principal characteristics of LAMP include the use of four to six primers annealing to initially four then six target sites selected to meet well established criteria, e.g., Eiken, 2018. The amplification mechanism is strand extension with loop formation producing what is often described as a cauliflower-like product, and a characteristic ladder band appearance on confirming gels. The multiple primer-target combination gives the process a very high degree of specificity, enabling target detection in crude preparations containing extraneous nucleic acids. The specificity also permits effective multiplex applications. The process uses a polymerase having strand extension activity, typically Bst, acting at constant temperature in the 60-70°C range. Operation at constant temperature permits amplification with simple means of maintaining constant temperature such as a water bath facilitating application in areas of limited laboratory facilities. The process has been found insensitive to interferences common to conventional PCR processes applied to analysis of environmental samples. Furthermore, the process is fully as sensitive as conventional PCR and amplification times, e.g. Ct, are typically short, i.e., 30-60 min, Broughton et al, 2020.

The relatively slow adoption of LAMP procedure for environmental monitoring specifically to water and wastewater is partly characteristic of processes having few detailed published reports to stimulate the interest of other investigators. Two features of LAMP may be described as disadvantages: 1) the rather intricate process of primer design, testing, and optimization needed to permit routine application; and 2) the very high sensitivity would permit cross contamination if not recognized and precluded by proper laboratory procedure and careful technique.

As a product of the world-wide spread of SARS-CoV-2 infection and COVID-19 disease throughout the human population has stimulated massive effort to develop and improve ability to detect and to monitor the virus e.g. Color, 2020. The standard method being applied at the beginning of the 4^th^ quarter of 2020 both to clinical detection and environmental monitoring, sewage, is PCR, typically RT qPCR, CDC, 2020; WHO, 2020. However, building on previous clinical applications many variations of LAMP-based procedure have been reported, Becherer et al, 2020. Not requiring a thermocycler, the process lends itself to both scale up and miniaturization, and can be combined with increasingly sophisticated technology and downstream refinements including sequencing.

The potential for successful application of LAMP to SARS-CoV-2 monitoring in raw sewage was a product of our previous experience applying a multiplex LAMP to detection of *Cryptosporidium* and *Giardia* in surface water samples (Ongerth and Saaed, 2020). In that work we found: 1) that a LAMP for each organism could be multiplexed for detection of both simultaneously; 2) the organisms were detectable at low concentration, 1-5 oocysts/cysts, concentrated from 10 L samples only by filtration and centrifuging to form a crude particle pellet subject to 10 freeze-thaw (f-t) cycles; 3) detection was not affected by extraneous components in the f-t processed pellet; and 4) quantification using a qPCR instrument (Roche Light Cycler 480) was possible. From early reports on monitoring SARS-CoV-2 in raw sewage, Ahmed et al, 2020: Wurtzer et al, 2020; Wu et al 2020, calculation of likely virus concentrations at a sewage treatment plant serving a population having COVID-19 daily reported cases in the range 5-10/100,000, suggested that the virus would b**e** detectable without concentration and that LAMP would not be affected by extraneous sewage components. To test the potential, taking advantage of many well-described LAMP primer sets reported for clinical application since April, 2020, Dong et al, 2020, we selecte**d** primers for three potential targets, ORF-1a (Lamb et al, 2020) E-gene and N-gene (Broughton et al, 2020), assembled essential materials, and arranged to obtain raw sewage samples with the local wastewater agency, East Bay Municipal Utility District Special District 1 (EBMUD SD1).

## Methods

Development, selection, and optimization of amplification conditions for each set of primers, ORF-1a, E-gene, and N-gene, was described in the original references, Lamb et al 2020, and Broughton et al, 2020. Primers were applied here using all of the concentration and amplification conditions established in the original references, (ORF1a, Lamb et al 2020; E-gene and N-gene, Broughton et al, 2020). To facilitate testing the approach we used off-the shelf materials where possible. Materials used included:

- Primers prepared with standard desalting, IDT, Coralville, IA
- Master mix: WarmStart LAMP Kit E1700, E2019, New England BioLabs, Ipswich, MA
- Control: Synthetic SARS-CoV-2 RNA, Control 6 (MT118835), Twist Bioscience, S. San Francisco, CA.
- Raw sewage: East Bay Municipal Utility District SD1 (EBMUD), Oakland, CA

Reactions for RT-qLAMP were prepared using 25 µL total volume, according to proportions listed in Table 2.

Upon receiving fresh sewage samples, reaction components were distributed into 0.2 mL PCR reaction tubes on ice during preparation. Reactions were prepared in a PCR hood using routine lab technique designed to preclude potential cross contamination. Amplifications were conducted using a Rotor-Gene Q programmed according to Qiagen protocol for constant temperature, 65°C (63°C after initial runs), for 30-40 minutes followed by high resolution melting. All reactions were conducted in triplicate. Reaction tubes were not opened after run completion and were frozen to permit future analysis.

Raw sewage was obtained from the EBMUD Special District 1 (SD1) serving a population of ca. 650,000 largely in Alameda County, Figure 2. Separate regions of the service are**a** contribute flows to the three interceptor sewers, North (N), Adeline (ADA), and South (S) terminating at the single treatment plant. Samples of 1L total volume were collected from each interceptor individually at a point just before the treatment plant. Samples were 24-hour composites representative of the period 9:00 am to 9:00 am, Figure 2c. Samples were transported on ice to the laboratory for processing and analysis, kept refrigerated w/o preservative until analysis. Sampling dates were July 29, Sept. 9, and Sept. 22.

**Figure 1.**
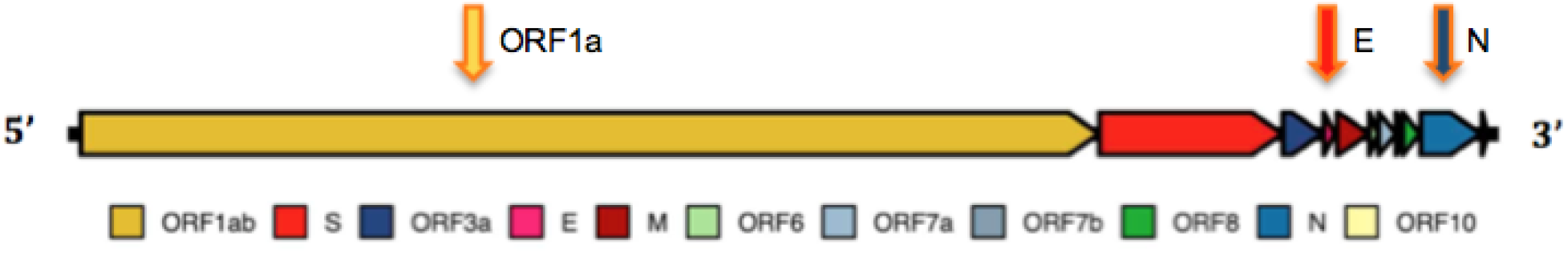
SARS-CoV-2 genome and subunit arrangement with primer locations

**Figure 2.**
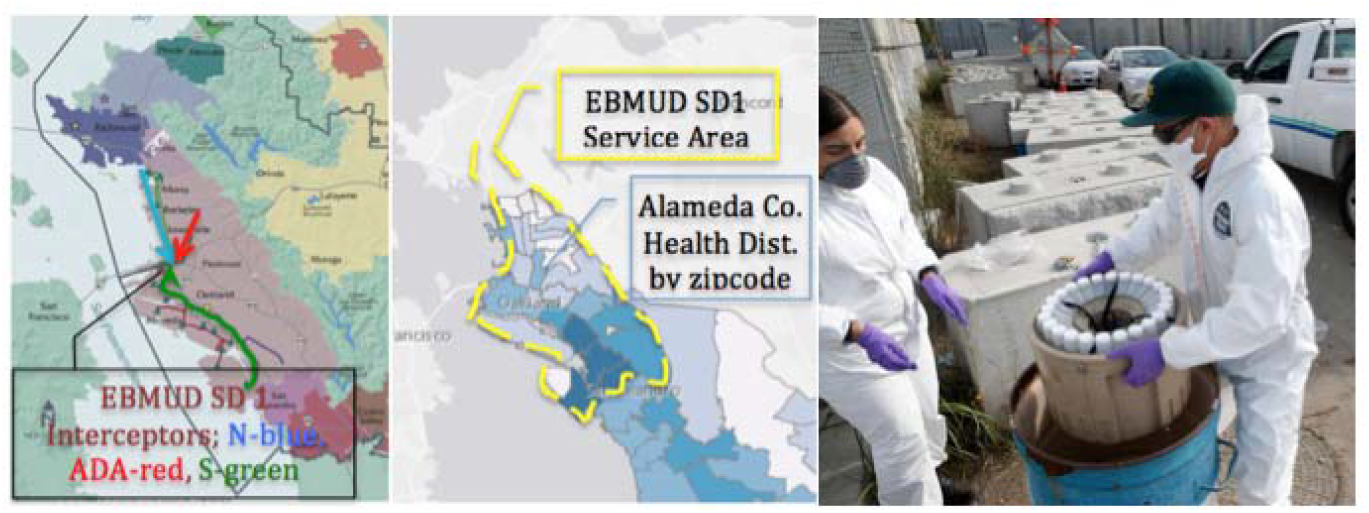
Left: EBMUD SD1 service area (mauve) with North (blue), Adaline (Red), and South (green) interceptors; Center: Alameda County Health District COVID-19 monitoring by zipcode; Right: EBMUD SD1 staff retrieving 24 hour samples for compositing.

## Results

Initial testing of RT LAMP to determine basic performance characteristics was begun using primers to the ORF1a gene. Testing consisted of 5-logs reference standard dilution from 10^4^-10^0^, including raw sewage. No acceptable standard curve could be obtained although raw sewage produced consistent amplification with Ct ca. 20-35, details below. Performance of RT qLAMP was then compared for E-gene and N-gene primers, Figure 3a-f. Standard curve quality was improved but remained low and significant differences in synthetic control amplification were apparent between E-gene and N-gene amplifications. We continued to include raw sewage with continued apparent amplification. Using melt curves as indicator, raw sewage SARS-CoV-2 RNA amplification appears specific. Not all replicates of either standards or sewage produced product, generally, standards at < 100 copies gave inconsistent reproducibility.

**Figure 3a-f.**
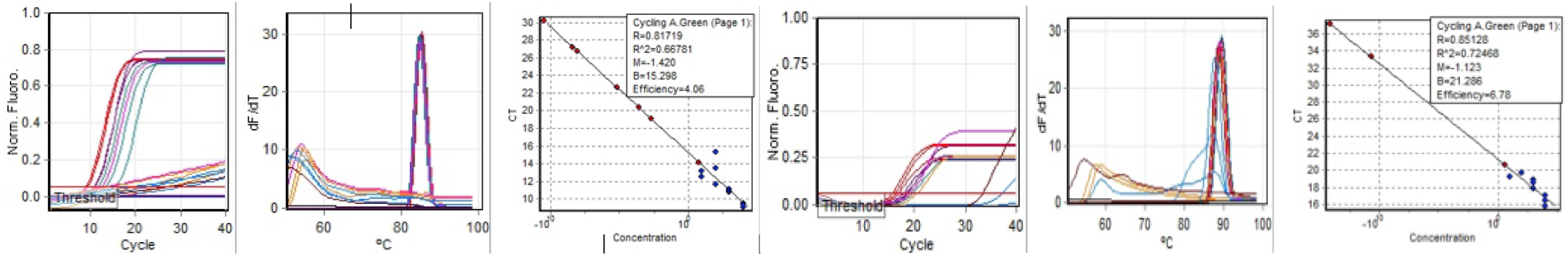
(left to right): E-gene amplification, melt, and standard curve; N-gene amplification, melt, and standard curve.

Further comparison of E-gene and N-gene performance was made examining the effect of reaction mix components, continuing to include raw sewage in amplification runs. Direct comparison between the previous run, September 9, without alteration of conditions was made on September 23. Standard curve quality for both E and N primers was improved. Amplification of synthetic RNA standards was less consistent although more consistent amplification from sewage was observed, Figures 4a-f.

**Figure 4a-f.**
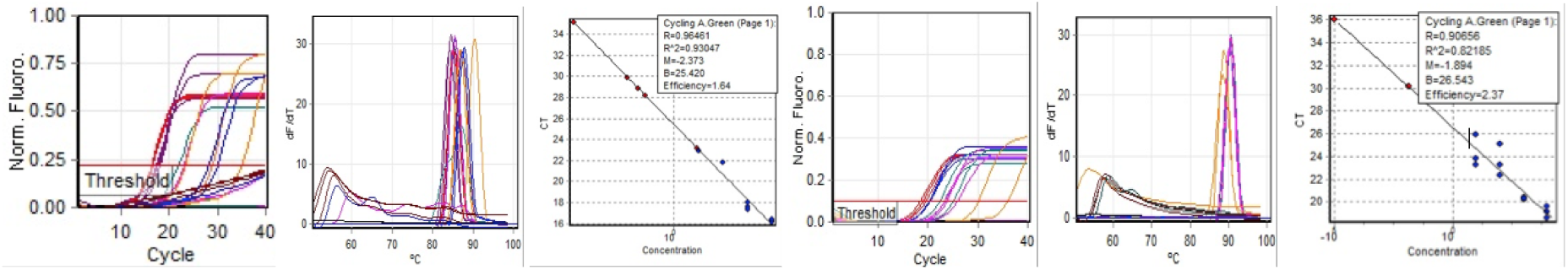
(left to right): E-gene amplification, melt, and standard curve; N-gene amplification, melt, and standard curve.

In companion runs on the same day, different mix components were used; NEB reaction mix 2019 was used instead of NEB reaction mix 1700.

As noted above, detection of SARS-CoV-2 in raw sewage was apparent throughout testing of RT LAMP and RT qLAMP performance. Analysis includes three sets of raw sewage samples from each of the three EBMUD SD1 interceptor sewers. Samples from July 29 produced only qualitative results due to inability to produce a standard curve using the ORF1a primers. Accordingly, all raw sewage amplification results are summarized in terms of Average Ct for each of the three sampling dates (7.29, 9.9, and 9.22), Figure 6a. Based on more acceptable performance resulting from amplifications summarized above, Figures 3, 4, and 5, quantitative product was determined based on 7μL raw sewage component of the 25 μL reaction mix, expressed as SARS-CoV-2/L, Figure 6b.

**Figure 5a-f.**
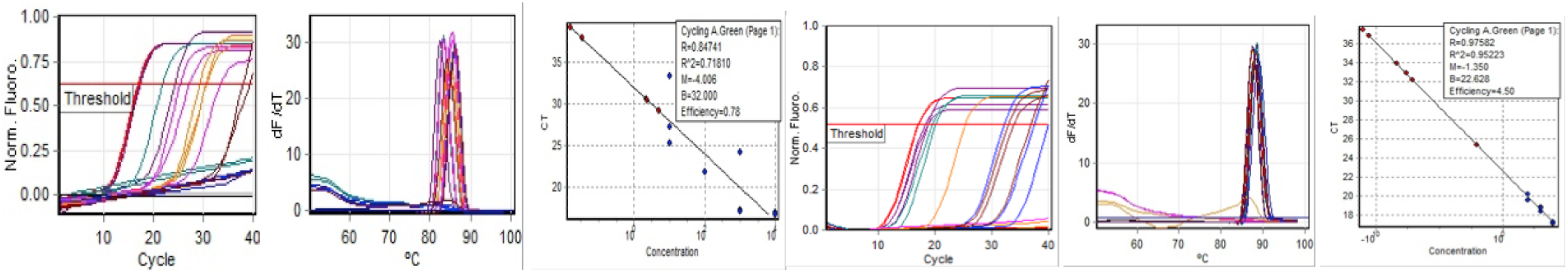
(left to right): E-gene amplification, melt, and standard curve; N-gene amplification, melt, and standard curve.

**Figure 6a.**
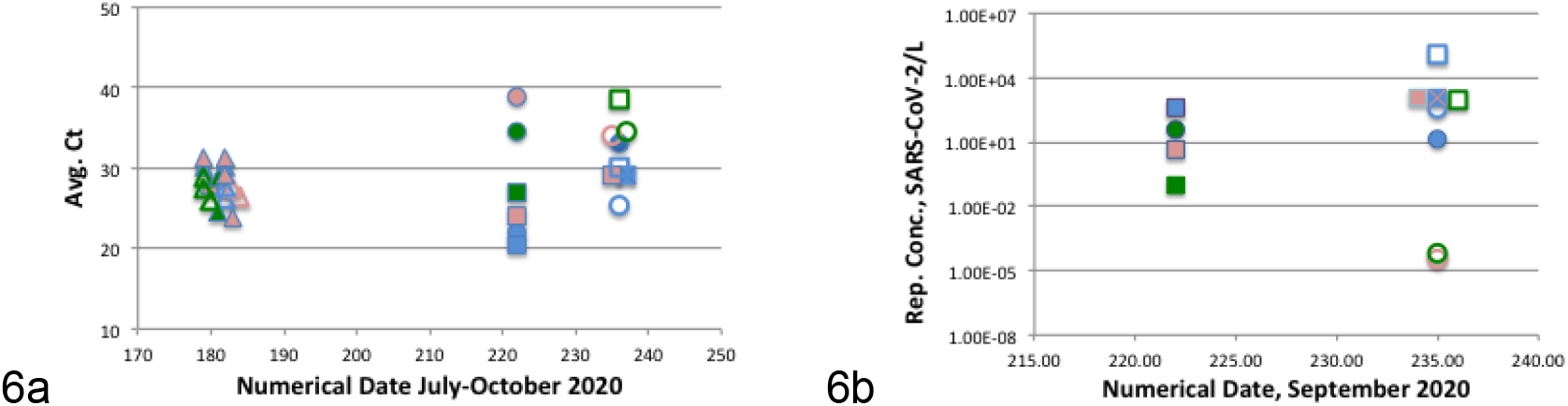
Raw sewage amplification product, Average Ct, samples July 29, September 9, and September 22. Raw sewage amplification product, SARS-CoV-2/L, samples September 9 and September 22. Symbol Key: **Site**: N=Blue; ADA=Red; S=Green; **Gene**: ORF1a=triangle, N=circle; E =square; **Mix**: 1700=filled; 2019=open.

## Discussion

Finding that RNA from SARS-CoV-2 in raw sewage can be amplified directly without pretreatment of concentration using RT LAMP may seem surprising. However, several carefully considered factors support the strength of the findings. First, the RT LAMP process has been widely and successfully applied to detection of SARS-CoV-2 in clinical samples beginning early this year, 2020, in response to COVID-19 monitoring needs, Thompson et al, 2020. The specific primers used in the work described here were selected from recent and thorough work to develop effective diagnostic tools. Second, accumulating information from both clinical assays and wastewater monitoring, e.g., Wu et al, July 2020, indicate that early COVID-19 infection, likely preceding onset of symptoms, results in fecal shedding bursts, estimated in the 10^12^/day range. Based on this level, shedding by a single individual in 100,000 would contribute to a SARS-CoV-2 concentration at the sewage treatment plant of ca. 2.5 x 10^5^/L or ca. 1.5 in a 5 μL template volume for a 25 μL LAMP reaction. And, third, LAMP as a process has been widely demonstrated to be sufficiently sensitive to amplify template at this level, i.e., 1-10 target copies/μL. Finally, and equally important, due to the multiple primer design and isothermal action polymerase, the LAMP process has selectivity permitting specific amplification in the presence of extraneous nucleic acids and other components of environmental media, sewage specifically that interfere with more common PCR analytical methods.

The work described here should be considered preliminary although providing clear evidence of SARS-CoV-2 detection. The poor performance of the ORF1a primers limiting quantification may have been partly due to its timing early in the sequence of testing. Although we made no procedural changes in succeeding runs, the quality of LAMP performance does appear to improve over the course of the six-week testing period. Continuing work is in progress to retest ORF1a performance.

Amplification characteristics observed differed between E-gene and N-gene primers, Figures 4 and 5. Initiation of amplification appeared somewhat earlier with E-gene primers and appreciably more product resulted. Variation in melt curve peaks was observed with E-gene amplifications less apparent in N-gene results. The peak melting temperature for N-gene product was 2-3° higher than for E-gene product.

The reactions in Figure 5 were conducted using the NEB E2019 reaction mix. It differs from the E1700 mix used in Figure 4 reactions by inclusion of dTTP, dUTP, and a thermolabile uracil DNA glycosylase (UDG). With consistent use in a sequence of amplification runs and incorporation of dU into amplification products, the presence of UDG will prevent potential carryover from previous reactions but will not affect amplification of the subsequent run due to complete inactivation at 65 °C. Our reactions had no predecessors using the E2019 mix so that effects observed would have been due to action of the additional mix components on amplification of both control synthetic RNA and components in the raw sewage. Whatever the mechanism, use of the mix increased product formation from both E and N gene primers, and appreciably improved the efficiency of N-gene reactions. It is important to note that although the NEB E2019 kit is supplied with E-gene and N-gene primers (both sets different from the E and N-gene primers used in our work (Table 1), we did not use the NEB primers. All our reactions used only the Table 1 primers.

**Table 1.**
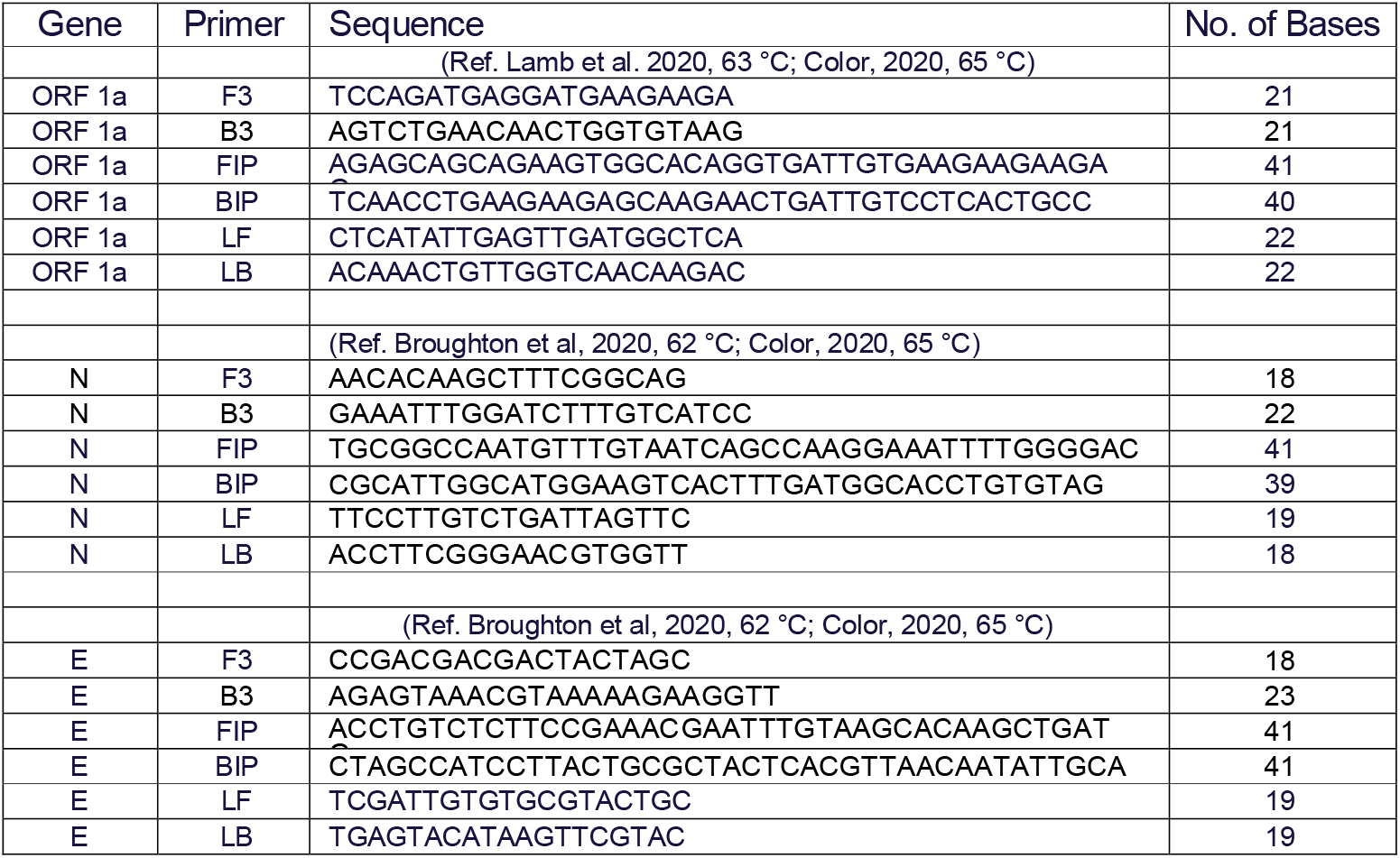
Primer sequences used for direct raw sewage RT qLAMP

**Table 2:** Direct raw sewage RT qLAMP reaction mix components

| Reaction Mix Component | Volume |
| --- | --- |
| WarmStart master mix (1700 or 2019) | 12.5 µL, includes dNTPs at 1.4 mM; RTx, 8 mM MgSO<sub>4</sub> |
| 10X Primers | 2.5 µL (1.6 µM FIP/BIP, 0.2 µM F3/B3, 0.4 µM Loop F/B) |
| NEB green fluorescent w/ E1700 kit | 0.5 µL (@ 0.5 µM) |
| Nuclease free H<sub>2</sub>O | 4.5 - 0 µL (adjust with template to 25 µL total rxn volume) |
| Template: Control or Raw sewage | 5 - 9.5 µL |
| Total Reaction Mix | 25 µL |

Ability to detect SARS-CoV-2 in sewage is fundamentally dependent on the extent of COVID-19 infection in the community. A reasonably detailed record of infection history in the EBMUD SD1 service area is maintained by the Alameda County Health Department (ACHD), ACHD, 2020. From the end of July through September corresponding to the period of our sewage analysis, the daily reported cases averaged ca. 150/day or for a population of just over 150,000, ca. 10/100,000 per day, Figure 7. The data show highly differentiated rates of infection among areas of the County and sewerage service area, cataloged according to zipcode.

**Figure 7.**
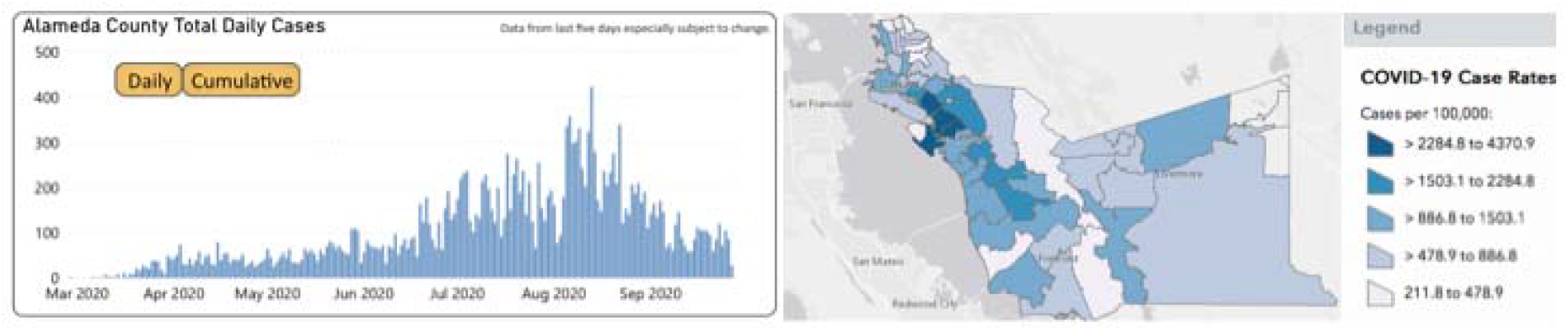
COVID-19 cases reported daily, March-September, 2020, and cumulative cases/100,000 by postal code, Alameda County, California.

The total cases reported, March 15 to September 30 (200 days) is 21,240, an average of 100 per day or 6.3/100,000 per day. Cumulative case rates among post code areas range from ca. 300/100,000 to nearly 2000/100,000. Understanding approximate incidence rates is important to the utility of monitoring sewage for understanding COVID-19 dynamics.

Considering how monitoring for SARS-CoV-2 in sewage may be of use in control of transmission several factors must be taken into account. These considerations have a direct bearing on the LAMP method described here and on needs for its refinement. A critical factor is the shedding rate in early, perhaps pre-symptomatic infections. Calculations described above indicate that detectable virus would be present in sewage for shedding at 10^12^ per day for a single infection/100,000 population. But, detection at this level is not a challenge to current detection methods. Many reports have shown that ample SARS-CoV-2 can be measured at the sewage treatment plant indicating only that infection is amply distributed in the community. To be useful for infection control the need is to be able to identify and if possible to isolate and trace contacts of the small number of early, high…perhaps super spreading…infections. Accordingly, monitoring focused on population concentrations of 100-1000 such as in institutions, hotels, multistory apartment building, industrial sites, will increase the sensitivity to detect by factors of 100 to 1000 in relation to the original assumption of 1 infection/100,000.

Recognizing that the real value of a method is ability to apply it to focused upstream sampling serves as a guide to features of the analytical method needing refinement and optimization. The most challenging problem of an RT qLAMP is refinement at minimal target concentration. While theoretically capable of amplification from a single copy…i.e. a single SARS-CoV-2 in a volume compatible with a 25 μL reaction mix for example, i.e. 5-10 μL, if the concentration in the sewage being sampled is that low, Poisson statistics dictate that a single copy will be present in only a minor proportion of replicated 5-10 μL aliquots analyzed. However, as suggested above, upstream sampling magnifies the concentration from a single shedder in direct proportion to the reduced population in the target sewage source. Thus, refinement of a RT qLAMP procedure for lowest limit of detection would not be important. Demonstration of consistently reproducible amplification at moderate concentrations is essential. Such demonstration should be readily achievable. Additional features require further attention including treatment of samples to make viral RNA available for amplification. We used freeze-thaw cycles and have no indication of adverse effects. The Mg^++^ concentration is a reaction mix component important to optimize. The primer-polymerase combination is sensitive to the total Mg^++^ concentration, optimized for our primers at 8 mM. Raw sewage includes Mg^++^; in our raw sewage samples ca. 33 mg/L. We did not adjust or re optimize to take this into account for the work described here. Future work must account for this component, likely to vary widely from among communities. Finally, the samples prepared by EBMUD SD1 staff were 24-hour composite samples. Samples collected at upstream sources are more likely to be grab samples and may be subject to variation over the typical 24-hour cycle of human activities that affect sewage composition around the clock.

Features of an RT qLAMP that are attractive for the type of application outlined above are those referenced in virtually all publications describing its advantages: LAMP is faster, cheaper, and more flexible than the RT qPCR procedure, currently the most widely applied method for sewage monitoring of SARS-CoV-2, CDC, 2020; WHO 2020. Sewage samples collected in the morning, returned to the lab can be combined with reaction mix components immediately and amplified…using the same thermocycler (or simple water bath) programmed for constant temperature…producing interpretable results in less than an hour. No preprocessing for virus concentration, RNA extraction, and interference mitigation is needed saving time, effort, and materials.

## Data Availability

All data contributing to this manuscript are available on request to the corresponding author.

## Acknowledgment

This work was made possible using the facilities of Cel Analytical Inc, San Francisco CA, ELAP# 2647, EPA ID CA 01529, and with the advice and consultation of Dr. Yeggi Dearborn, Cel Analytical owner.

